## Supplementary Material for "Generalizability Assessment of AI Models Across Hospitals: A Comparative Study in Low-Middle Income and High Income Countries"

#### A Software Packages and Implementation

Experiments were performed using Python (v3.6.9). All models were run using an Intel Xeon E-2146G Processor (CPU: 6 cores, 4.50 GHz max frequency). Logistic regression was implemented using Scikit-learn (v1.2.1). The XGBoost classifier was implemented using the XGBoost package (v1.3.0). Neural network models were implemented using Pytorch (1.13.1+cu117).

#### B Datasets

##### B.1 CURIAL Datasets

United Kingdom National Health Service (NHS) approval via the national oversight/regulatory body, the Health Research Authority (HRA), has been granted for development and validation of artificial intelligence models to detect Covid-19 (CURIAL; NHS HRA IRAS ID: 281832).

Data from OUH studied here are available from the Infections in Oxfordshire Research Database (https://oxfordbrc.nihr.ac.uk/research-themes/modernising-medical-microbiology-and-big-infection-diagnostics/infections-in-oxfordshire-research-database-iord/), subject to an application meeting the ethical and governance requirements of the Database. Data from UHB, PUH and BH are available on reasonable request to the respective trusts, subject to HRA requirements.

The following inclusions and exclusions are reproduced from previous studies (Soltan et al., 2022, Yang et al., 2022a, Yang et al., 2022b).

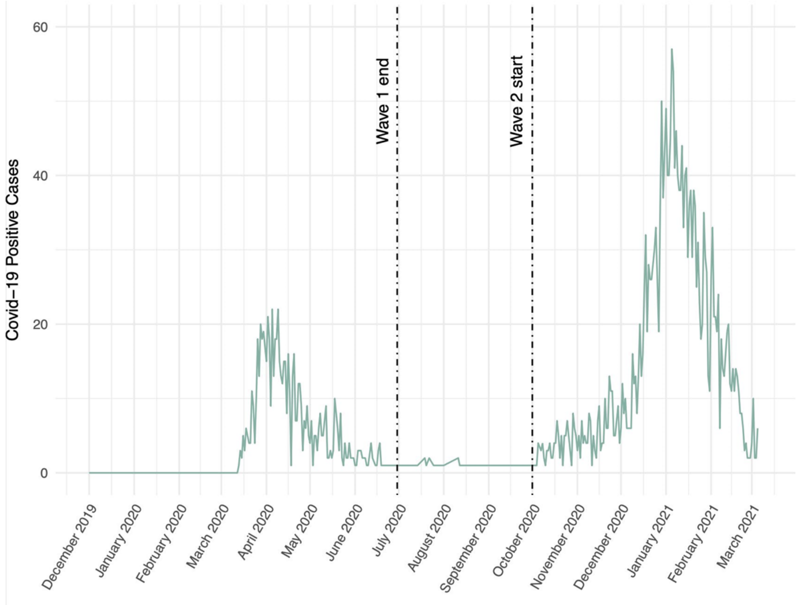


Supplementary Figure 1: Daily number of patients presenting to Oxford University Hospitals NHS Foundation Trust testing positive for COVID-19, between 1st December 2019 and 6th March 2021.

**Oxford University Hospitals NHS Foundation Trust (OUH):** We included all patients attending acute and emergency care settings at OUH who received routine blood tests on arrival, considering presentations before December 1, 2019, and thus before the pandemic, as the COVID-19-negative (control) cohort. We considered presentations during the 'first wave' of the UK COVID-19 pandemic (December 1, 2019 to June 30, 2020) with PCR confirmed SARS-CoV-2 infection as the COVID-19-positive (cases) cohort. We excluded patients who opted out of electronic health record (EHR) research and those who did not receive laboratory blood tests or were younger than 18 years of age. Due to incomplete penetrance of testing during the first wave of the pandemic, and imperfect sensitivity of the PCR test, there is uncertainty in the viral status of patients presenting during the pandemic who were untested or tested negative. We therefore selected a pre-pandemic control cohort during training to ensure absence of disease in patients labelled as COVID-19-negative. Clinical features extracted for each presentation included first-performed blood tests, blood gases, vital signs measurements and PCR testing for SARS-CoV-2 (Abbott Architect [Abbott, Maidenhead, UK], TaqPath [Thermo Fisher Scientific, Massachusetts, USA] and Public Health England-designed RNA-dependent RNA polymerase assays).

**Portsmouth Hospitals University NHS Foundation Trust (PUH):** PUH considered all patients admitted to the Queen Alexandria Hospital, serving a population of 675,000 and offering tertiary referral services to the surrounding region, between March 1, 2020 and February 28, 2021. Confirmatory COVID-19 testing was by laboratory SARS-CoV2 RT-PCR assay, considering any positive PCR result within 48hrs of admission as a true positive.

**University Hospitals Birmingham NHS Foundation Trust (UHB):** UHB considered all patients admitted to The Queen Elizabeth Hospital, Birmingham, between December 01, 2019 and October 29, 2020. The Queen Elizabeth Hospital is a large tertiary referral unit within the UHB group which provides healthcare services for a population of 2.2 million across the West Midlands. Confirmatory COVID-19 testing was performed by laboratory SARS-CoV-2 RT-PCR assay.

**Bedfordshire NHS Foundation Trust (BH):** BH considered all patients admitted to Bedford Hospital between January 1, 2021 and March 31, 2021. BH provides healthcare services for a population of around 620,000 in Bedfordshire. Confirmatory COVID-19 testing was performed on the day of admission by point-of-care PCR based nucleic acid testing [SAMBA-II & Panther Fusion System, Diagnostics in the Real World, UK, and Hologic, USA].

##### B.2 Vietnam Datasets

The ethics committees of the Hospital for Tropical Diseases (HTD) and the National Hospital for Tropical Diseases (NHTD) approved use of the HTD and NHTD datasets for COVID-19 diagnosis, respectively.

**Hospital for Tropical Diseases (HTD):** HTD considered all patients admitted between January 1, 2021 and December 31,2022.

**National Hospital for Tropical Diseases (NHTD):** NHTD considered all patients admitted between January 1, 2021 and December 31,2022.

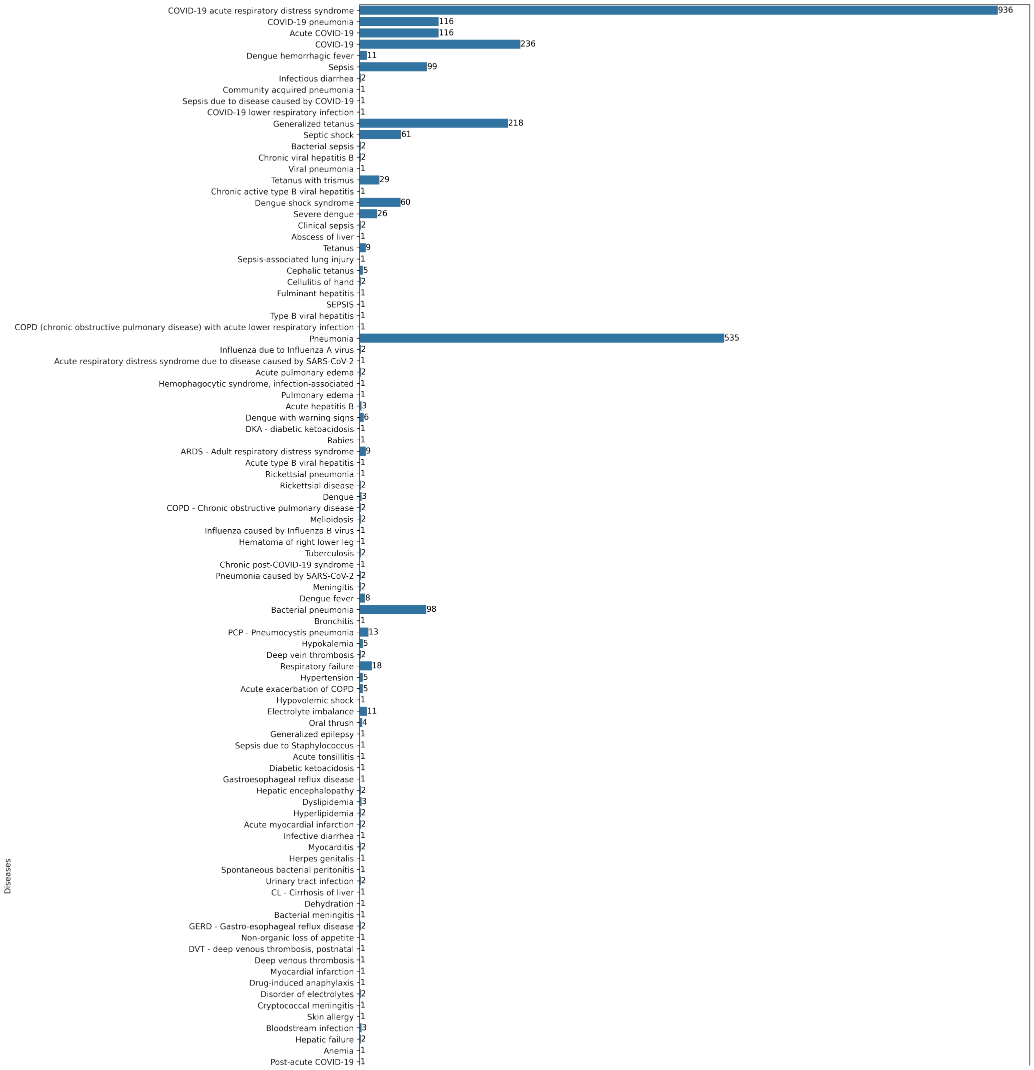


Supplementary Figure 2: Diseases recorded for patients admitted to the Hospital for Tropical Diseases. It should be noted that each patient's EHR data had two columns for listing disorders, and thus, the counts of all diseases is greater than the total number of patients in the cohort. Figure continues on next page.

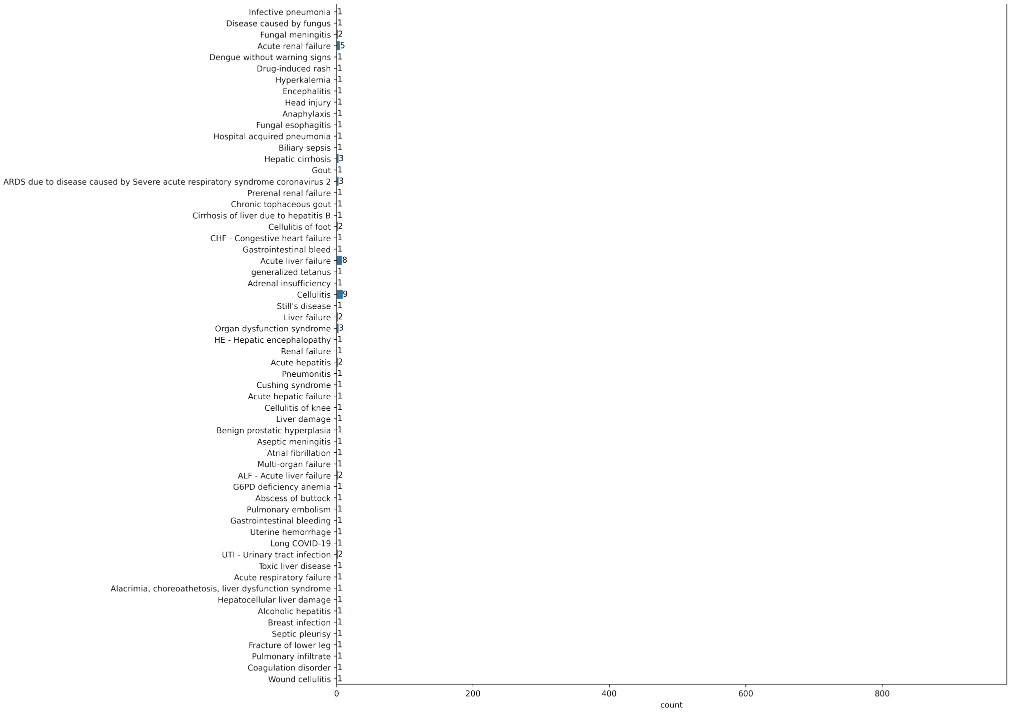
Infective pneumonia 1
Disease caused by fungus 1
Fungal meningitis 2
Acute renal failure 5
Dengue without warning signs 1
Drug-induced rash 1
Hyperkalemia 1
Encephalitis 1
Head injury 1
Anaphylaxis 1
Fungal esophagitis 1
Hospital acquired pneumonia 1
Biliary sepsis 1
Hepatic cirrhosis 3
Gout 1
ARDS due to disease caused by Severe acute respiratory syndrome coronavirus 2 3
Prerenal renal failure 1
Chronic tophaceous gout 1
Cirrhosis of liver due to hepatitis B 1
Cellulitis of foot 2
CHF - Congestive heart failure 1
Gastrointestinal bleed 1
Acute liver failure 8
generalized tetanus 1
Adrenal insufficiency 1
Cellulitis 9
Still's disease 1
Liver failure 2
Organ dysfunction syndrome 3
HE - Hepatic encephalopathy 1
Renal failure 1
Acute hepatitis 2
Pneumonitis 1
Cushing syndrome 1
Acute hepatic failure 1
Cellulitis of knee 1
Liver damage 1
Benign prostatic hyperplasia 1
Aseptic meningitis 1
Atrial fibrillation 1
Multi-organ failure 1
ALF - Acute liver failure 2
G6PD deficiency anemia 1
Abscess of buttock 1
Pulmonary embolism 1
Gastrointestinal bleeding 1
Uterine hemorrhage 1
Long COVID-19 1
UTI - Urinary tract infection 2
Toxic liver disease 1
Acute respiratory failure 1
Alacrimia, choreoathetosis, liver dysfunction syndrome 1
Hepatocellular liver damage 1
Alcoholic hepatitis 1
Breast infection 1
Septic pleurisy 1
Fracture of lower leg 1
Pulmonary infiltrate 1
Coagulation disorder 1
Wound cellulitis 1
0
200
400
600
800
count

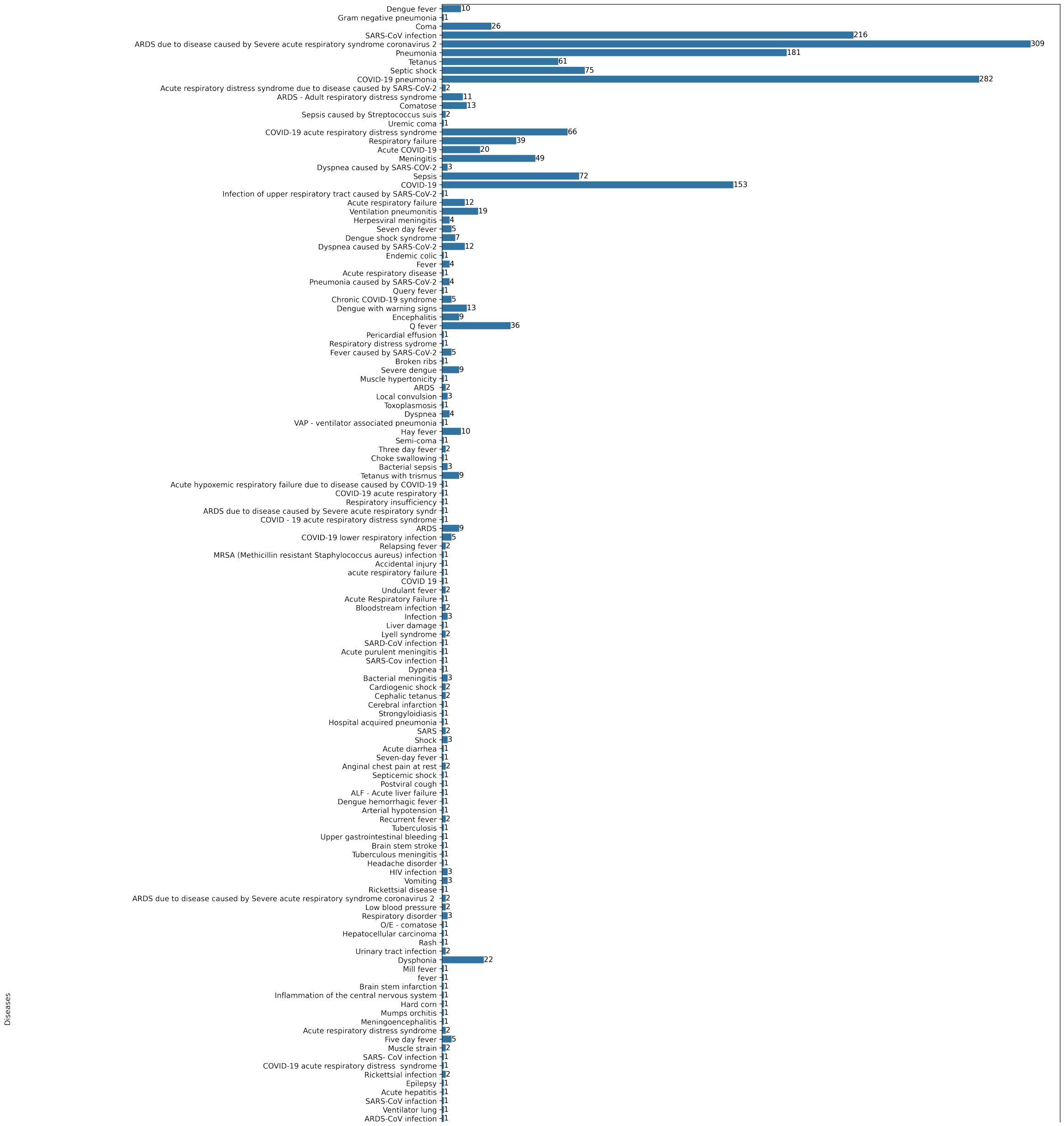


Supplementary Figure 3: Diseases recorded for patients admitted to the National Hospital for Tropical Diseases. It should be noted that each patient's EHR data had two columns for listing disorders, and thus, the counts of all diseases is greater than the total number of patients in the cohort. Figure continues on next page.

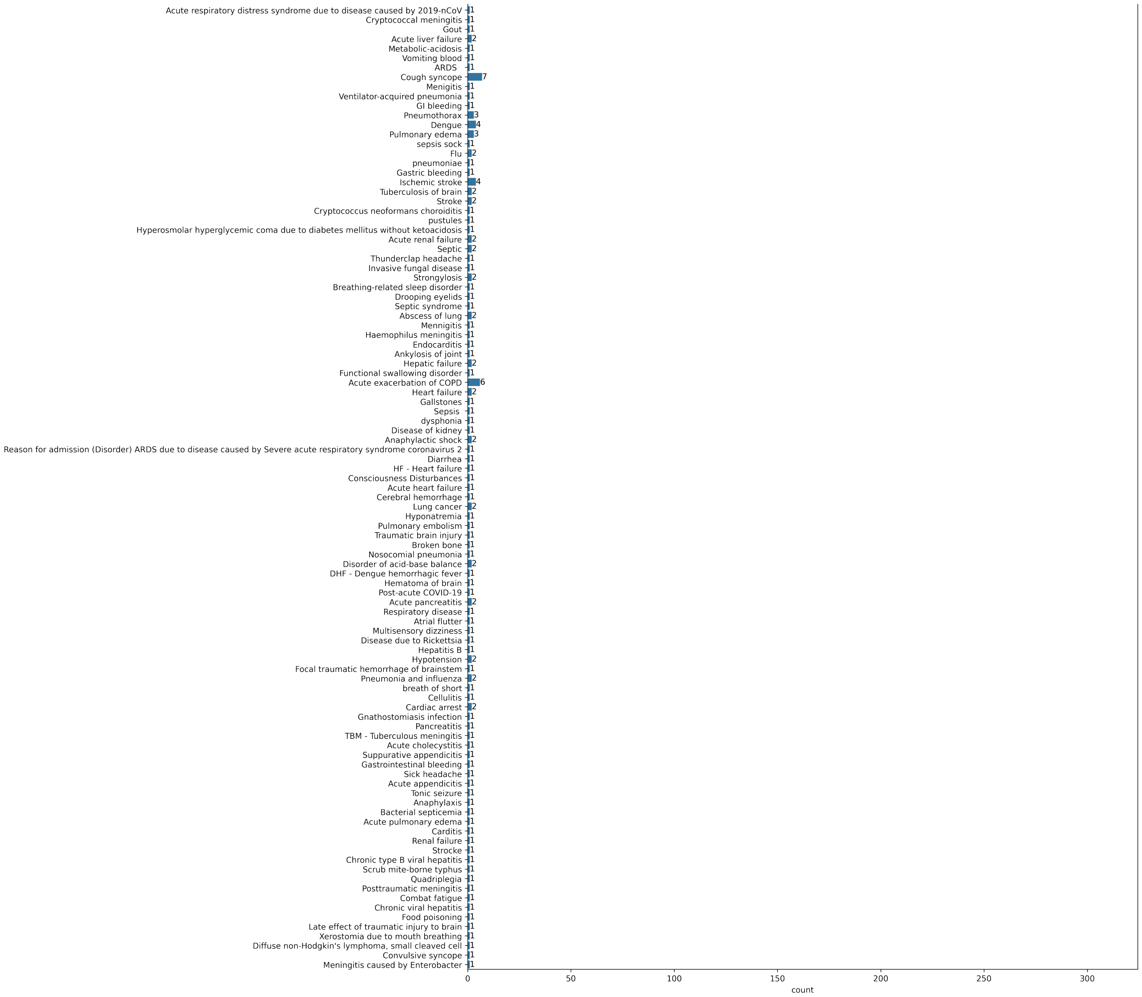


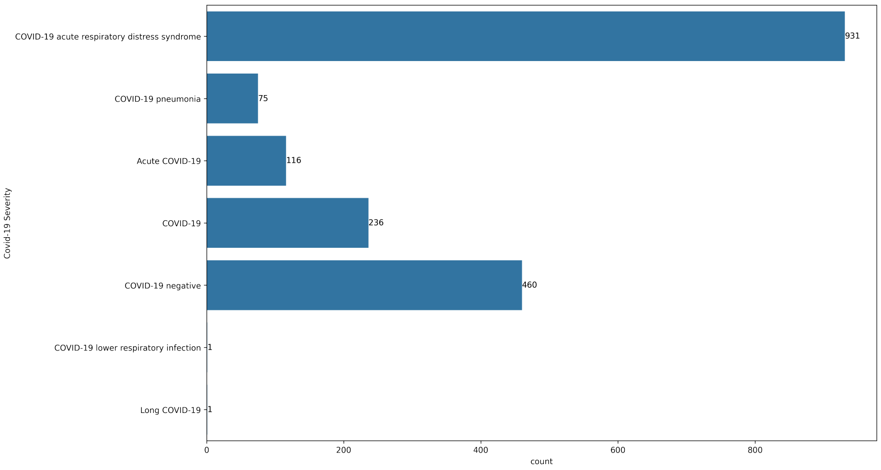


Supplementary Figure 4: Levels of COVID-19 severity recorded for patients admitted to the Hospital for Tropical Diseases.

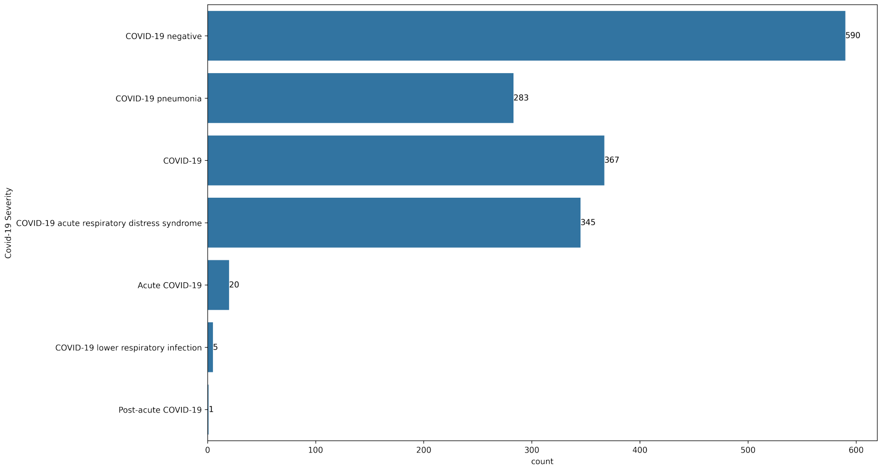


Supplementary Figure 5: Levels of COVID-19 severity recorded for patients admitted to the National Hospital for Tropical Diseases.

Supplementary Table 1: Distribution of vital signs, reported as median and interquartile ranges, for each patient cohort.

| | Oxford University Hospitals (pre-pandemic & wave 1 cases, to June 30/20) | | Oxford University Hospitals | Portsmouth Hospitals University NHS Trust | University Hospitals Birmingham NHS Foundation Trust | Bedfordshire Hospitals NHS Foundation Trust | Hospital for Tropical Diseases | National Hospital for Tropical Diseases | Kruskal-Wallis, p-value |
|---|---|---|---|---|---|---|---|---|---|
| | Prepandemic | COVID-19-cases | Oct 1/20-Mar 6/21 | Mar 1/20-Feb 28/21 | Dec 1/19 - Oct 29/20 | Jan 1/21-Mar 31/21 | Jan 1/21-Dec 31/22 | Jan 1/21-Dec 31/22 | |
| Respiratory Rate (breath/min) | 18.0 (16.0-19.0) | 20.0 (18.0-24.0) | 18.0 (16.6-19.0) | 17.0 (16.0-19.0) | 18.0 (17.0-20.0) | 18.0 (16.0-20.0) | 24.0(20.0-26.5) | 25.0(22.0-30.0) | <0.0001 |
| Heart Rate (beats/min) | 82.0 (71.0-96.0) | 88.0 (75.0-101.0) | 84.0 (72.0-97.0) | 82.0 (71.0-95.0) | 86.0 (73.0-101.0) | 84.0 (73.0-97.0) | 94.0(84.0-108.0) | 99.0(86.0-114.0) | <0.0001 |
| Systolic Blood Pressure (mmHg) | 132.0 (118.0-150.0) | 131.0 (115.0-146.0) | 134.0 (119.0-152.0) | 128.0 (114.0-146.0) | 136.0 (119.0-155.0) | 131.0 (116.0-149.0) | 130.0(111.5-140.0) | 120.0(110.0-136.0) | <0.0001 |
| Diastolic Blood Pressure (mmHg) | 74.0 (65.0-84.0) | 74.0 (64.0-84.0) | 75.0 (65.0-85.0) | 76.0 (67.0-84.0) | 77.0 (68.0-87.0) | 78.0 (68.0-88.0) | 80.0(70.0-80.0) | 70.0(60.0-80.0) | <0.0001 |
| Tympanic Temperature (C) | 36.5 (36.1-36.9) | 36.9 (36.3-37.6) | 36.3 (36.0-36.7) | 36.3 (36.0-36.8) | 36.7 (36.4-37.2) | 36.5 (36.4-36.9) | 37.0(37.0-37.3) | 37.0(36.8-37.5) | <0.0001 |

Supplementary Table 2: Distribution of blood test features, reported as median and interquartile ranges, for each patient cohort.

| | Oxford University Hospitals (pre-pandemic & wave 1 cases, to June 30/20) | | Oxford University Hospitals | Portsmouth Hospitals University NHS Trust | University Hospitals Birmingham NHS Foundation Trust | Bedfordshire Hospitals NHS Foundation Trust | Hospital for Tropical Diseases | National Hospital for Tropical Diseases | Kruskal-Wallis, p-value |
|---|---|---|---|---|---|---|---|---|---|
| | Prepandemic cohort | COVID-19-cases cohort | Oct 1/20-Mar 6/21 | Mar 1/20-Feb 28/21 | Dec 1/19 - Oct 29/20 | Jan 1/21-Mar 31/21 | Jan 1/21-Dec 31/22 | Jan 1/21-Dec 31/22 | |
| HAEMOGLOBIN (g/L) | 130.0 (116.0-142.0) | 130.0 (114.0-144.0) | 129.0 (114.0-142.0) | 129.0 (114.0-143.0) | 127.0 (113.0-140.0) | 134.0 (119.0-146.0) | 128.0(113.0-141.0) | 112.0(95.0-127.0) | <0.0001 |
| WHITE CELLS ($10^9 1^{-1}$) | 8.45 (6.46-11.18) | 6.98 (5.14-9.72) | 8.94 (6.7-12.06) | 8.6 (6.7-11.3) | 9.4 (7.1-12.6) | 9.2 (6.9-12.5) | 9.55(6.69-13.565) | 10.9(7.525-15.0) | <0.0001 |
| PLATELETS ($10^9 1^{-1}$) | 249.0 (199.0-307.0) | 215.0 (163.0-283.5) | 251.0 (198.0-314.0) | 251.0 (199.0-312.0) | 247.0 (196.0-311.0) | 246.0 (196.0-310.0) | 216.0(152.0-281.0) | 189.5(116.75-260.25) | <0.0001 |
| HAEMATOCRIT | 0.39 (0.35-0.42) | 0.4 (0.35-0.44) | 0.39 (0.35-0.43) | 0.39 (0.34-0.42) | 0.38 (0.34-0.42) | 0.39 (0.35-0.43) | 0.392(0.35-0.428) | 0.342(0.289-0.387) | <0.0001 |
| SODIUM (mM) | 138.0 (136.0-140.0) | 136.0 (134.0-139.0) | 138.0 (135.0-140.0) | 138.0 (136.0-140.0) | 137.0 (134.0-139.0) | 138.0 (136.0-140.0) | 134.0(130.0-137.0) | 136.0(132.65-139.65) | <0.0001 |
| UREA (mM) | 5.3 (4.0-7.4) | 5.9 (4.2-9.07) | 5.7 (4.2-8.3) | 5.2 (3.8-7.6) | 6.2 (4.5-9.0) | 5.8 (4.2-8.3) | 7.5(5.0-12.75) | 7.215(4.8-11.7) | <0.0001 |
| BILIRUBIN (umol/L) | 9.0 (6.0-13.0) | 9.0 (7.0-13.25) | 9.0 (6.0-14.0) | 10.0 (7.0-16.0) | 10.0 (7.0-15.0) | 10.0 (7.0-14.0) | 13.7(7.8-36.7) | 9.8(6.4-16.375) | <0.0001 |
| CREATININE (umol/L) | 73.0 (60.0-93.0) | 79.0 (65.0-106.0) | 74.0 (60.0-97.0) | 74.0 (60.0-96.0) | 78.0 (62.0-105.0) | 80.5 (65.75-104.0) | 76.0(61.0-99.0) | 77.65(56.0-121.0) | <0.0001 |
| POTASSIUM (mM) | 4.0 (3.7-4.3) | 4.0 (3.7-4.3) | 4.0 (3.8-4.4) | 4.2 (3.9-4.4) | 4.1 (3.8-4.4) | 4.3 (4.0-4.6) | 3.72(3.35-4.09) | 3.8(3.4-4.3) | <0.0001 |
| MEAN CELL VOL (fl) | 89.6 (86.0-93.4) | 90.2 (86.6-94.2) | 90.2 (86.6-94.2) | 89.0 (84.9-93.0) | 89.9 (86.2-93.6) | 88.0 (85.0-92.0) | | | 0.210 |
| NEUTROPHILS ($10^9 1^{-1}$) | 5.72 (3.99-8.36) | 5.11 (3.48-7.49) | 6.44 (4.4-9.55) | 5.9 (4.2-8.6) | 6.9 (4.7-10.0) | 6.8 (4.7-9.73) | | | <0.0001 |
| LYMPHOCYTES ($10^9 1^{-1}$) | 1.51 (1.0-2.13) | 0.96 (0.65-1.38) | 1.31 (0.85-1.89) | 1.5 (0.97-2.2) | 1.3 (0.9-1.9) | 1.27 (0.86-1.83) | | | <0.0001 |
| MONOCYTES ($10^9 1^{-1}$) | 0.64 (0.48-0.85) | 0.49 (0.35-0.74) | 0.66 (0.48-0.89) | 0.63 (0.48-0.85) | 0.7 (0.5-0.9) | 0.66 (0.48-0.92) | | | <0.0001 |
| EOSINOPHILS ($10^9 1^{-1}$) | 0.1 (0.04-0.2) | 0.01 (0.0-0.06) | 0.07 (0.02-0.16) | 0.1 (0.02-0.2) | 0.1 (0.0-0.2) | 0.06 (0.02-0.16) | | | <0.0001 |
| BASOPHILS ($10^9 1^{-1}$) | 0.04 (0.03-0.06) | 0.02 (0.01-0.03) | 0.04 (0.02-0.06) | 0.04 (0.02-0.06) | 0.1 (0.0-0.1) | 0.05 (0.03-0.07) | | | <0.0001 |
| ALBUMIN (g/L) | 36.0 (32.0-39.0) | 32.0 (28.0-35.0) | 36.0 (31.0-39.0) | 36.0 (31.0-40.0) | 36.0 (32.0-40.0) | 35.0 (31.0-39.0) | | | 0.006 |
| ALKALINE PHOSPHATASE (IU/L) | 80.0 (64.0-105.0) | 82.0 (64.0-108.0) | 84.0 (66.0-112.0) | 84.0 (67.0-109.0) | 90.0 (71.0-119.0) | 94.0 (74.5-122.0) | | | <0.0001 |
| ALT (IU/L) | 18.0 (13.0-28.0) | 25.0 (17.0-41.0) | 20.0 (13.0-33.0) | 19.0 (13.0-30.0) | 19.0 (13.0-30.0) | 20.0 (13.0-31.0) | | | <0.0001 |
| eGFR (ml/min) | 85.0 (63.0-150.0) | 78.0 (53.0-150.0) | 84.0 (58.0-150.0) | 83.0 (60.0-90.0) | 76.0 (52.0-90.0) | 76.0 (54.0-90.0) | | | 0.011 |
| CRP (mg/L) | 8.6 (2.3-39.0) | 72.5 (23.8-143.6) | 15.8 (3.5-67.4) | 13.0 (3.0-71.0) | 12.0 (3.0-61.0) | 10.7 (2.8-48.78) | | | 0.003 |

### C Model Architectures

**Neural Network Models**: The rectified linear unit (ReLU) activation function was used for the hidden layers and the Sigmoid activation function was used in the output layer. For updating model weights, binary cross entropy loss and the Adaptive Moment Estimation (Adam) optimizer was used during training. Models were trained using 100 epochs and a batch size of 2048. Add summary of model

```
CovidClassifier(
  (hidden1): Linear(in_features=14, out_features=10, bias=True)
  (act1): ReLU()
  (output): Linear(in_features=10, out_features=1, bias=True)
  (act_output): Sigmoid()
)
```

Supplementary Figure 6: Neural network architecture used in COVID-19 classification task.

### D Results

Supplementary Table 3: COVID-19 diagnosis performance across logistic regression, XGBoost, and neural network models trained on the UK data. Results are presented for the reduced feature set. Results are optimized to sensitivities of 0.85. Bolded values denote the best scores across models for each test set. Metrics are reported alongside 95% confidence intervals (CIs).

| Test Set | Model | AUROC | AUPRC | Sensitivity | Specificity | PPV | NPV |
|---|---|---|---|---|---|---|---|
| OUH | LR | 0.795(0.785-0.805) | 0.35(0.334-0.37) | 0.692(0.676-0.709) | **0.79(0.785-0.794)** | **0.241(0.235-0.247)** | **0.964(0.962-0.966)** |
| | XGB | 0.784(0.774-0.793) | 0.408(0.388-0.426) | 0.711(0.693-0.727) | 0.716(0.711-0.721) | 0.195(0.19-0.199) | 0.962(0.96-0.964) |
| | NN | **0.803(0.793-0.812)** | **0.428(0.408-0.448)** | **0.718(0.701-0.733)** | 0.725(0.72-0.73) | 0.201(0.197-0.206) | **0.964(0.962-0.966)** |
| PUH | LR | 0.812(0.803-0.821) | 0.222(0.209-0.235) | 0.781(0.765-0.796) | 0.696(0.692-0.7) | 0.126(0.123-0.128) | 0.983(0.981-0.984) |
| | XGB | **0.817(0.808-0.825)** | **0.346(0.326-0.365)** | 0.766(0.75-0.781) | **0.709(0.704-0.712)** | **0.128(0.125-0.131)** | 0.982(0.981-0.983) |
| | NN | 0.816(0.808-0.825) | 0.307(0.29-0.326) | **0.835(0.822-0.849)** | 0.592(0.587-0.596) | 0.103(0.101-0.104) | **0.985(0.983-0.986)** |
| UHB | LR | 0.757(0.735-0.782) | 0.196(0.169-0.228) | 0.599(0.562-0.64) | **0.821(0.815-0.827)** | **0.129(0.122-0.138)** | 0.979(0.977-0.981) |
| | XGB | 0.76(0.74-0.78) | 0.191(0.163-0.225) | 0.617(0.58-0.656) | 0.754(0.747-0.762) | 0.101(0.095-0.107) | 0.978(0.976-0.98) |
| | NN | **0.776(0.755-0.797)** | **0.242(0.21-0.279)** | **0.69(0.654-0.726)** | 0.743(0.736-0.75) | 0.107(0.102-0.113) | **0.982(0.98-0.984)** |
| BH | LR | 0.801(0.768-0.833) | 0.363(0.308-0.433) | 0.549(0.48-0.616) | **0.861(0.842-0.878)** | **0.354(0.314-0.395)** | 0.932(0.922-0.941) |
| | XGB | 0.773(0.735-0.807) | 0.409(0.344-0.481) | 0.576(0.508-0.646) | 0.811(0.791-0.831) | 0.299(0.265-0.332) | 0.932(0.922-0.943) |
| | NN | **0.804(0.772-0.838)** | **0.442(0.377-0.519)** | **0.688(0.619-0.754)** | 0.74(0.717-0.763) | 0.269(0.244-0.296) | **0.944(0.933-0.956)** |
| HTD | LR | 0.488(0.465-0.515) | 0.717(0.694-0.74) | 0.896(0.883-0.91) | 0.1(0.077-0.124) | 0.746(0.741-0.752) | 0.246(0.196-0.295) |
| | XGB | 0.533(0.508-0.559) | 0.755(0.732-0.779) | 0.803(0.786-0.82) | **0.202(0.172-0.236)** | 0.748(0.741-0.757) | 0.258(0.227-0.293) |
| | NN | **0.577(0.551-0.604)** | **0.763(0.741-0.786)** | **0.908(0.895-0.921)** | 0.139(0.114-0.167) | **0.757(0.751-0.764)** | **0.339(0.287-0.395)** |
| NHTD | LR | 0.471(0.446-0.497) | 0.631(0.606-0.66) | 0.824(0.805-0.843) | 0.161(0.136-0.187) | 0.65(0.641-0.659) | 0.327(0.284-0.372) |
| | XGB | 0.478(0.453-0.504) | 0.633(0.608-0.663) | 0.727(0.703-0.75) | **0.272(0.242-0.304)** | **0.654(0.641-0.666)** | **0.346(0.313-0.379)** |
| | NN | **0.515(0.491-0.541)** | **0.671(0.647-0.7)** | **0.831(0.812-0.85)** | 0.159(0.134-0.185) | 0.651(0.642-0.66) | 0.333(0.288-0.377) |

Supplementary Table 4: COVID-19 diagnosis performance across logistic regression, XGBoost, and neural network models trained on the UK data. Results are presented for the comprehensive (GATS-filled) dataset. Results are optimized to sensitivities of 0.85. Bolded values denote the best scores across models for each test set. Metrics are reported alongside 95% confidence intervals (CIs).

| Test Set | Model | AUROC | AUPRC | Sensitivity | Specificity | PPV | NPV |
|---|---|---|---|---|---|---|---|
| OUH | LR | 0.854(0.845-0.862) | 0.472(0.452-0.493) | 0.738(0.721-0.754) | **0.85(0.846-0.854)** | **0.322(0.314-0.329)** | 0.971(0.969-0.973) |
| | XGB | **0.877(0.869-0.885)** | **0.636(0.618-0.654)** | **0.815(0.799-0.829)** | 0.774(0.769-0.778) | 0.258(0.252-0.263) | **0.977(0.976-0.979)** |
| | NN | 0.865(0.856-0.872) | 0.587(0.568-0.606) | 0.77(0.753-0.785) | 0.8(0.795-0.804) | 0.271(0.265-0.277) | 0.973(0.971-0.975) |
| PUH | LR | 0.832(0.823-0.84) | 0.308(0.292-0.325) | 0.759(0.744-0.774) | **0.753(0.75-0.757)** | 0.147(0.144-0.15) | 0.982(0.981-0.984) |
| | XGB | **0.877(0.869-0.884)** | **0.506(0.486-0.525)** | **0.825(0.81-0.839)** | 0.748(0.744-0.751) | **0.154(0.151-0.158)** | **0.987(0.986-0.988)** |
| | NN | 0.853(0.844-0.861) | 0.433(0.414-0.454) | 0.786(0.771-0.801) | **0.753(0.75-0.757)** | 0.151(0.148-0.154) | 0.984(0.983-0.985) |
| UHB | LR | 0.846(0.828-0.864) | 0.297(0.264-0.34) | 0.747(0.712-0.782) | **0.832(0.826-0.839)** | **0.166(0.158-0.174)** | 0.987(0.985-0.988) |
| | XGB | **0.86(0.845-0.876)** | 0.38(0.342-0.422) | **0.779(0.745-0.812)** | 0.787(0.781-0.794) | 0.14(0.134-0.147) | **0.988(0.986-0.989)** |
| | NN | 0.859(0.841-0.876) | **0.386(0.347-0.431)** | 0.772(0.737-0.803) | 0.795(0.788-0.802) | 0.144(0.137-0.151) | 0.987(0.985-0.989) |
| BH | LR | 0.883(0.856-0.908) | 0.519(0.454-0.597) | 0.66(0.594-0.721) | **0.918(0.904-0.932)** | **0.528(0.482-0.58)** | 0.951(0.942-0.959) |
| | XGB | **0.905(0.881-0.926)** | **0.669(0.607-0.729)** | **0.792(0.732-0.848)** | 0.849(0.83-0.867) | 0.422(0.39-0.46) | **0.967(0.958-0.976)** |
| | NN | 0.875(0.846-0.902) | 0.579(0.51-0.654) | 0.688(0.623-0.747) | 0.873(0.856-0.891) | 0.43(0.391-0.472) | 0.952(0.943-0.961) |
| HTD | LR | 0.492(0.466-0.519) | 0.728(0.706-0.752) | 0.571(0.549-0.593) | 0.435(0.396-0.474) | 0.749(0.735-0.764) | 0.255(0.236-0.276) |
| | XGB | 0.539(0.514-0.566) | 0.774(0.752-0.798) | 0.646(0.625-0.668) | 0.404(0.367-0.44) | 0.762(0.749-0.774) | 0.278(0.256-0.299) |
| | NN | **0.59(0.563-0.617)** | **0.784(0.762-0.808)** | **0.661(0.64-0.682)** | **0.465(0.426-0.505)** | **0.785(0.772-0.799)** | **0.317(0.295-0.34)** |
| NHTD | LR | 0.475(0.449-0.498) | 0.641(0.616-0.671) | 0.54(0.516-0.564) | **0.432(0.397-0.465)** | 0.642(0.624-0.658) | 0.332(0.311-0.352) |
| | XGB | 0.484(0.458-0.508) | 0.642(0.617-0.671) | 0.61(0.585-0.633) | 0.369(0.335-0.404) | 0.646(0.632-0.662) | 0.334(0.31-0.359) |
| | NN | **0.522(0.497-0.544)** | **0.68(0.655-0.707)** | **0.66(0.635-0.682)** | 0.353(0.319-0.385) | **0.658(0.644-0.672)** | **0.355(0.328-0.382)** |

Supplementary Table 5: COVID-19 diagnosis performance across neural network models, trained using transfer learning at HTD and NHTD centres. Models are evaluated prospectively on the site used for transfer learning (remaining subset not used in training) and externally on the other site (full dataset). Results are presented for both the reduced feature set, and the comprehensive feature set. Sensitivity is optimized to 0.85. Metrics are reported alongside 95% confidence intervals (CIs).

| Features | Training Set | Test Set | AUROC | AUPRC | Sensitivity | Specificity | PPV | NPV |
|---|---|---|---|---|---|---|---|---|
| Reduced | HTD | HTD | 0.707(0.654-0.756) | 0.769(0.715-0.828) | 0.876(0.841-0.911) | 0.418(0.342-0.491) | 0.749(0.725-0.775) | 0.63(0.557-0.706) |
| | | NHTD | 0.653(0.627-0.677) | 0.748(0.723-0.776) | 0.855(0.837-0.874) | 0.328(0.294-0.361) | 0.706(0.695-0.718) | 0.545(0.505-0.586) |
| | NHTD | HTD | 0.656(0.599-0.712) | 0.818(0.77-0.869) | 0.816(0.773-0.855) | 0.386(0.3-0.471) | 0.78(0.755-0.804) | 0.442(0.365-0.52) |
| | | NHTD | 0.65(0.623-0.675) | 0.815(0.795-0.84) | 0.801(0.785-0.819) | 0.422(0.385-0.46) | 0.804(0.793-0.815) | 0.418(0.388-0.448) |
| Full | HTD | HTD | 0.727(0.676-0.774) | 0.792(0.745-0.845) | 0.897(0.863-0.926) | 0.467(0.391-0.541) | 0.77(0.744-0.795) | 0.695(0.621-0.77) |
| | | NHTD | 0.663(0.639-0.686) | 0.758(0.732-0.785) | 0.877(0.861-0.895) | 0.269(0.236-0.299) | 0.694(0.684-0.703) | 0.538(0.497-0.583) |
| | NHTD | HTD | 0.668(0.61-0.722) | 0.836(0.794-0.877) | 0.769(0.721-0.813) | 0.489(0.402-0.575) | 0.8(0.772-0.829) | 0.443(0.378-0.513) |
| | | NHTD | 0.69(0.666-0.713) | 0.845(0.826-0.868) | 0.747(0.728-0.766) | 0.524(0.486-0.56) | 0.823(0.811-0.835) | 0.412(0.387-0.438) |

Supplementary Table 6: COVID-19 diagnosis performance using locally-trained models. Results are presented for neural network models, trained using a subset of data from either the HTD or NHTD centres. Models are evaluated prospectively on the site used for model development (i.e. remaining subset not used in training) and externally on the other site (full dataset). Sensitivity is optimized to 0.85. Metrics are reported alongside 95% confidence intervals (CIs).

| Training Set | Test Set | AUROC | AUPRC | Sensitivity | Specificity | PPV | NPV |
|---|---|---|---|---|---|---|---|
| HTD | HTD | 0.664(0.613-0.716) | 0.75(0.699-0.806) | 0.868(0.833-0.904) | 0.369(0.296-0.436) | 0.732(0.709-0.755) | 0.584(0.506-0.664) |
| | NHTD | 0.639(0.615-0.663) | 0.739(0.714-0.768) | 0.849(0.831-0.868) | 0.305(0.271-0.338) | 0.697(0.687-0.709) | 0.517(0.476-0.559) |
| NHTD | HTD | 0.608(0.585-0.634) | 0.808(0.788-0.832) | 0.76(0.741-0.78) | 0.378(0.342-0.415) | 0.783(0.773-0.794) | 0.348(0.322-0.377) |
| | NHTD | 0.662(0.604-0.717) | 0.829(0.784-0.873) | 0.786(0.742-0.83) | 0.455(0.366-0.542) | 0.793(0.766-0.822) | 0.444(0.374-0.515) |
